## Supplementary Information for "Cross-organ Analysis Reveals Associations between Vascular Properties of the Retina, the Carotid and Aortic Artery, and the Brain"

#### Data

Supplementary Data 1 - Data IDPs information [1]

Supplementary Data 2 - Data Main IDPs information [2]

Supplementary Data 3 - Phenotypic correlations [3]

Supplementary Data 4 - Genetic corrleations [4]

Supplementary Data 5 - Heritabilities [5]

Supplementary Data 6 - Data shared genes [6]

Supplementary Data 7 - Data shared pathways [7]

Supplementary Data 8 - Data genes coherence [8]

Supplementary Data 9 - Temporal analysis [9]

---

[1] https://docs.google.com/spreadsheets/d/1zX7WXg7ioAj2Z0iEbhTrRIMnYWuylARDSMebdV-Rh7o/edit?pli=1#gid=0

[2] https://docs.google.com/spreadsheets/d/1zX7WXg7ioAj2Z0iEbhTrRIMnYWuylARDSMebdV-Rh7o/edit?pli=1#gid=1617304176

[3] https://docs.google.com/spreadsheets/d/1zX7WXg7ioAj2Z0iEbhTrRIMnYWuylARDSMebdV-Rh7o/edit?pli=1&gid=555028989#gid=555028989

[4] https://docs.google.com/spreadsheets/d/1zX7WXg7ioAj2Z0iEbhTrRIMnYWuylARDSMebdV-Rh7o/edit?pli=1&gid=607908070#gid=607908070

[5] https://docs.google.com/spreadsheets/d/1zX7WXg7ioAj2Z0iEbhTrRIMnYWuylARDSMebdV-Rh7o/edit?pli=1&gid=44958827#gid=44958827

[6] https://docs.google.com/spreadsheets/d/1zX7WXg7ioAj2Z0iEbhTrRIMnYWuylARDSMebdV-Rh7o/edit?pli=1#gid=1387648808

[7] https://docs.google.com/spreadsheets/d/1zX7WXg7ioAj2Z0iEbhTrRIMnYWuylARDSMebdV-Rh7o/edit?pli=1#gid=448785807

[8] https://docs.google.com/spreadsheets/d/1zX7WXg7ioAj2Z0iEbhTrRIMnYWuylARDSMebdV-Rh7o/edit?gid=1909207584#gid=1909207584

[9] https://docs.google.com/spreadsheets/d/1zX7WXg7ioAj2Z0iEbhTrRIMnYWuylARDSMebdV-Rh7o/edit?pli=1&gid=370245248#gid=370245248

### 1 Main Vascular IDPs Selection

For vascular image-derived phenotypes (IDPs) with over 1,000 participants, we analyzed both or morphological and functional IDPs. In the brain, we had access to cerebral blood flow (CBF) and arterial transit time (ATT) across various regions. To simplify the analysis while capturing how functional IDPs relate to morphological ones, we plotted the phenotypic correlation, after adjusting for covariates (Supplementary Figure 1). This figure illustrates the high correlations among these IDPs. Consequently, we averaged these measures to obtain general CBF and ATT values for the brain.

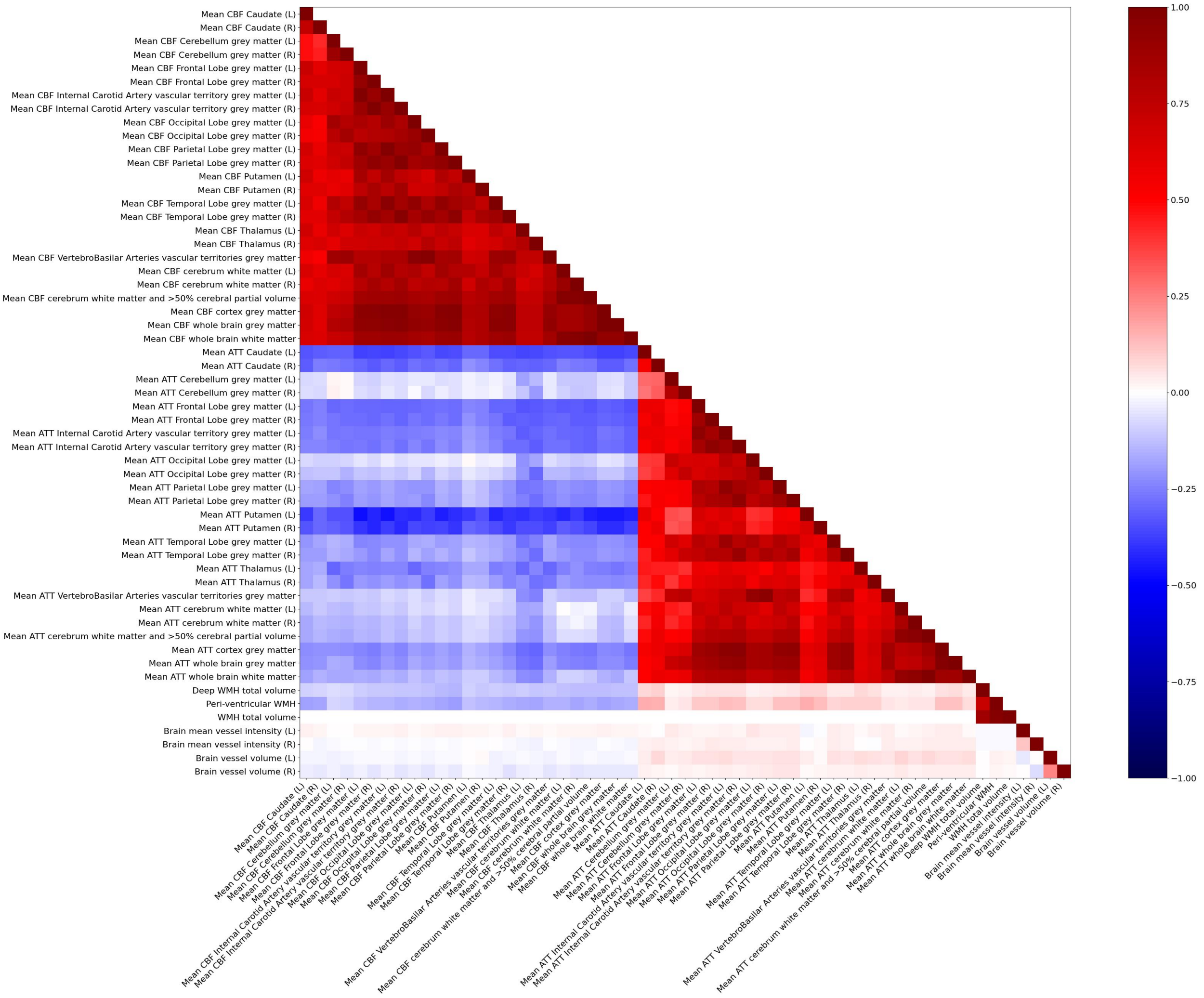


Figure 1: Phenotypic correlation of brain IDPs. This figure shows the correlation between geometric and functional brain IDPs, after adjusting for covariates and with outliers beyond ± 10 std removed. All IDPs were adjusted for covariates (see Methods in the main document). Abbreviations: R: right hemisphere, L: left hemisphere.

After reducing the list of IDPs by averaging CBF and ATT, we assessed the phenotypic associations of morphological and functional non-retinal IDPs with each other, as well as with retinal IDPs (Supplementary Figure 2). This analysis helps to understand how functional IDPs relate to morphological ones. This figure reveals that some morphological IDPs, such as minimum (min), maximum (max), and mean intima-media thickness (IMT), are highly correlated. Thus, we retained only the min IMT as a main IDP, and similarly simplified the list of aortic areas IDPs.

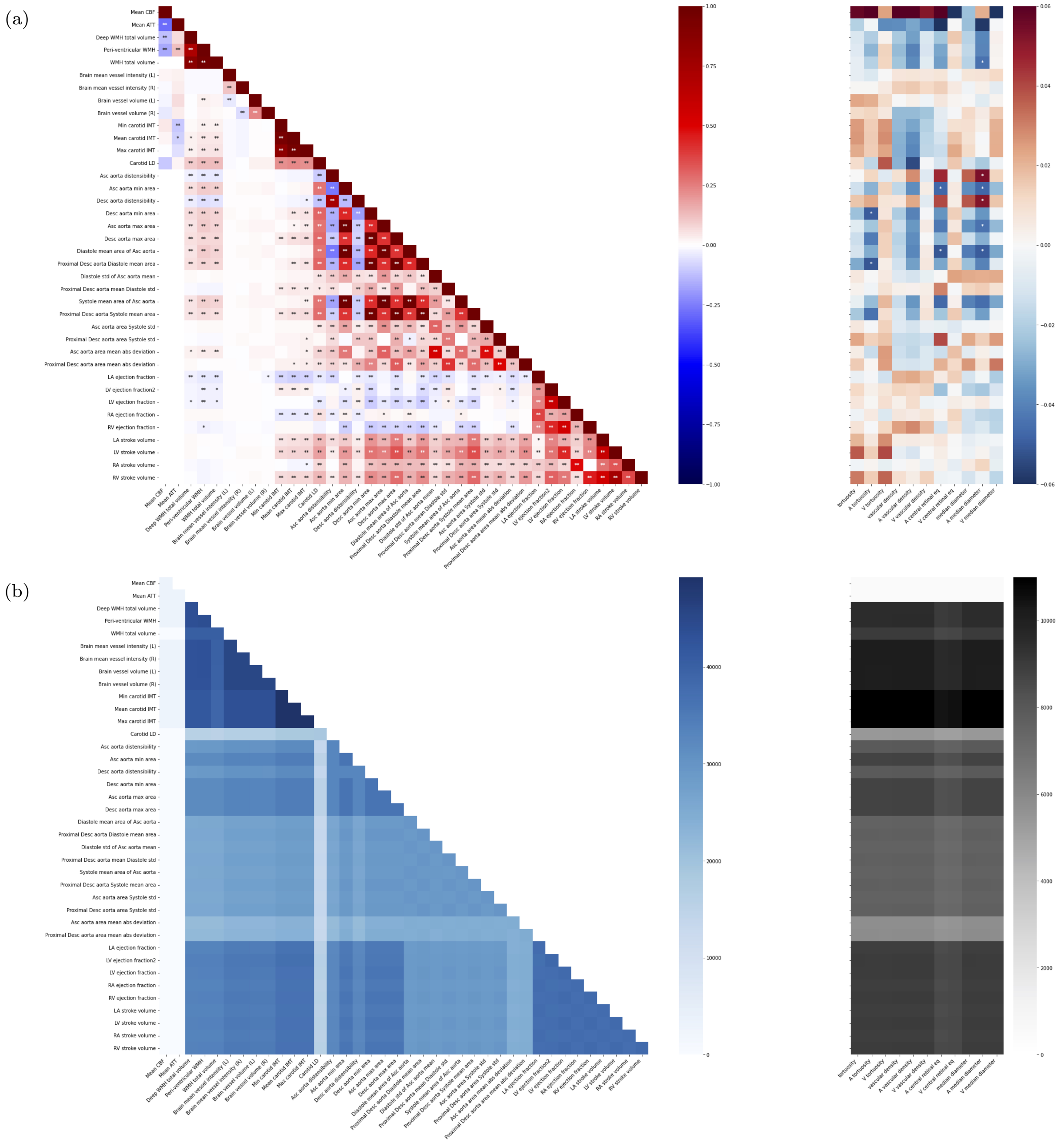

Figure 2: **a)** Phenotypic association of morphological and functional non-retinal IDPs with non-retinal IDPs (**left**) and retinal IDPs (**right**). The x-axis represents non-retinal IDPs (**left**) and retinal IDPs (**right**), while the y-axis shows non-retinal IDPs. Colors indicate standardized effect sizes for linear regressions, and asterisks denote statistical significance with p-values corrected for multiple testing ($* : p < 0.05/N_{test}$, $** : p < 0.001/N_{test}$, where $N_{test} = N_{IDPs} \times (N_{IDPs}/2 + N_{retina})$. IDPs after adjusting for covariates (see Methods in the main document). **b)** Number of subjects per IDP pair. Abbreviations: LA: left atrium, LV: left ventricle, RA: right atrium, RV: right ventricle.

#### 2 Number of Subjects Main IDPs

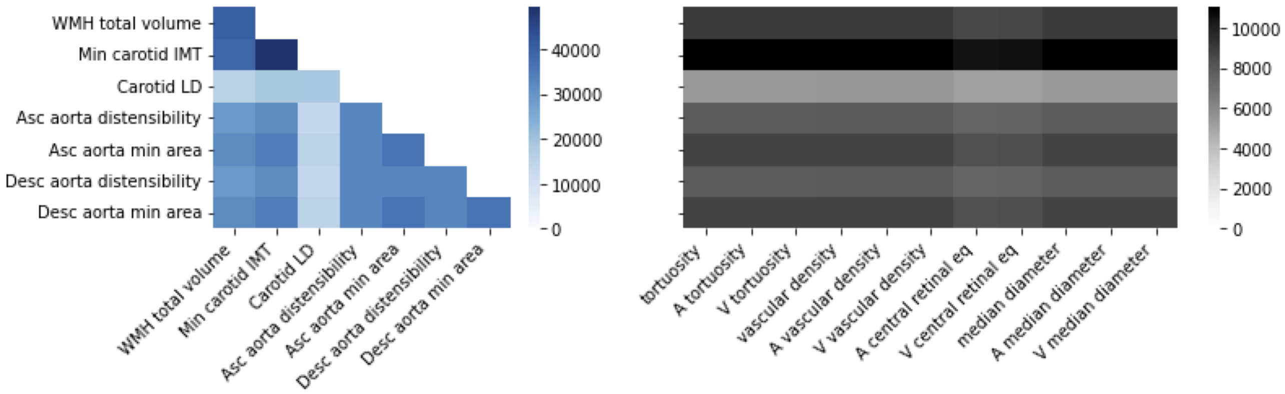


Figure 3: Number of subjects per pair of main IDPs. This figure corresponds to the figures of the main document 2.a and 2.b.

#### 3 Sensitivity Analyses with Extended Cardiovascular Risk Adjustment

To assess the robustness of the IDP correlations to additional cardiovascular risk factors, we performed sensitivity analyses in which we incrementally expanded the covariate set used in the main analyses (Figure 2b). In addition to the demographic and blood pressure covariates already included (age, age$^2$, sex, 10 genetic principal components, and hypertension status), we further adjusted for: (i) diabetes status, (ii) smoking status, and (iii) high-density lipoprotein (HDL) cholesterol level. These variables were derived from UK Biobank (UKB) fields: '2443' (diabetes diagnosis), '20116' (smoking status), and '30760' (HDL cholesterol).

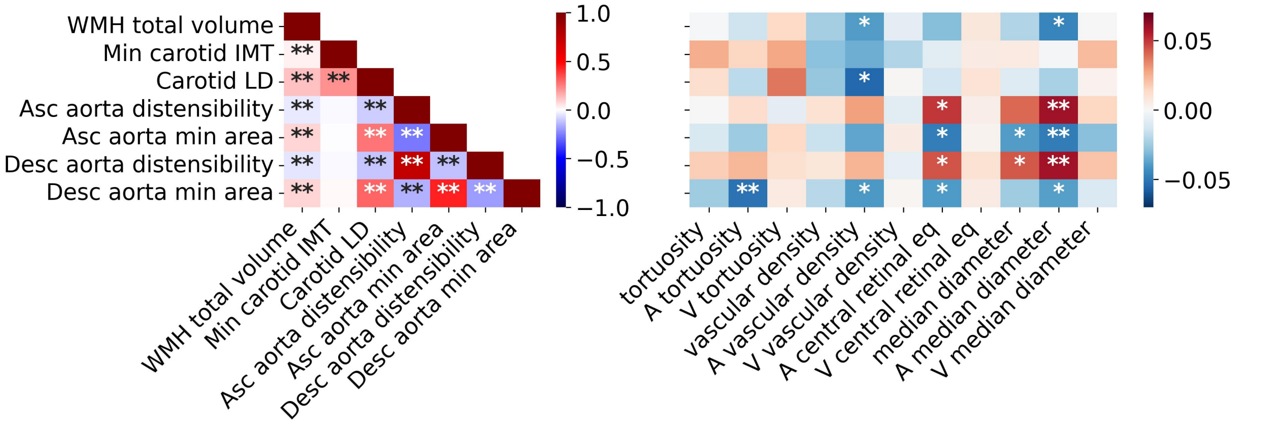


Figure 4: Extended cardiovascular risk adjustment sensitivity analyses. Phenotypic correlation of non-retinal IDPs with non-retinal IDPs (**left**) and retinal IDPs (**right**), additionally adjusted for hypertension, diabetes, smoking, and HDL. Hypertension was defined as having a systolic blood pressure (SBP) ≥140 mmHg or diastolic blood pressure (DBP) ≥90 mmHg.

#### 4 Sensitivity to Imaging–Acquisition Date

In the primary analysis, we combined retinal imaging-derived phenotypes (IDPs) captured during UKB instances 0–1 (2006–2013) with non-retinal IDPs—carotid ultrasound and MRI—recorded at instance 2 (2014+). The median time gap between modalities is therefore ∼9 years, raising the concern that secular or age-related trends might spuriously inflate or attenuate cross-organ correlations.

For that reason, we repeated the phenotypic-correlation analysis after dividing the retinal sample into two non-overlapping sub-cohorts:

- **Reduced-gap cohort**: participants whose retinal images were acquired at instance 1 (2012–2013) and who possessed at least one, non-retinal IDP acquired within $\leq 2$ years of the fundus examination ($N = 19{,}354$).
- **Long-gap cohort**: participants whose retinal images came from instance 0 (2006–2010), providing an upper bound on the acquisition gap.

For each retinal–non-retinal pair, we retained all individuals common to both modalities within the specified cohort (the reduced-gap and long-gap). Down-sampling the long-gap cohort to mirror the smaller cohort would have reduced the pair IDP sample size much more than the reduced-gap cohort. Therefore, we analysed each cohort at its intrinsic size, which led to more similar sample sizes per IDP pairs (Supplementary Figures 5b and 5d). All correlations were computed after regressing out the same covariates as in the main Figure 2a and applying $z$-scaling within modality.

The principal cross-organ associations were almost identical across the two cohorts in both direction and magnitude (Supplementary Figures 5a and 5c). For example, white-matter hyperintensity (WMH) burden remained positively correlated with retinal arteriolar vessel density ($r = -0.04$, in the main figure (which combines both instances); $r = -0.06$, in the reduced-gap cohort; $r = -0.04$, in the long-gap cohort). A similar preservation was observed for carotid lumen diameter (LD) ($r = -0.06$, in the main figure; $r = -0.06$, in the reduced-gap cohort; $r = -0.07$, in the long-gap cohort) and ascending aorta distensibility ($r = 0.06$ in the main figure; $r = 0.07$, in the reduced-gap cohort; $r = 0.05$, in the long-gap cohort). Collectively, these results indicate that the temporal gap does not distort the cross-organ correlation structure.

Supplementary Figure 5 shows the full correlation matrices (panels a, c) and their corresponding IDP-pair sample-sizes (panels b, d). Bonferroni-significant threshold was applied ($n = 77$ tests; *: p<0.05/n=0.000649 and **: p<0.01/n=0.000130). For more details, see Supplementary Data 9 - Temporal analysis.

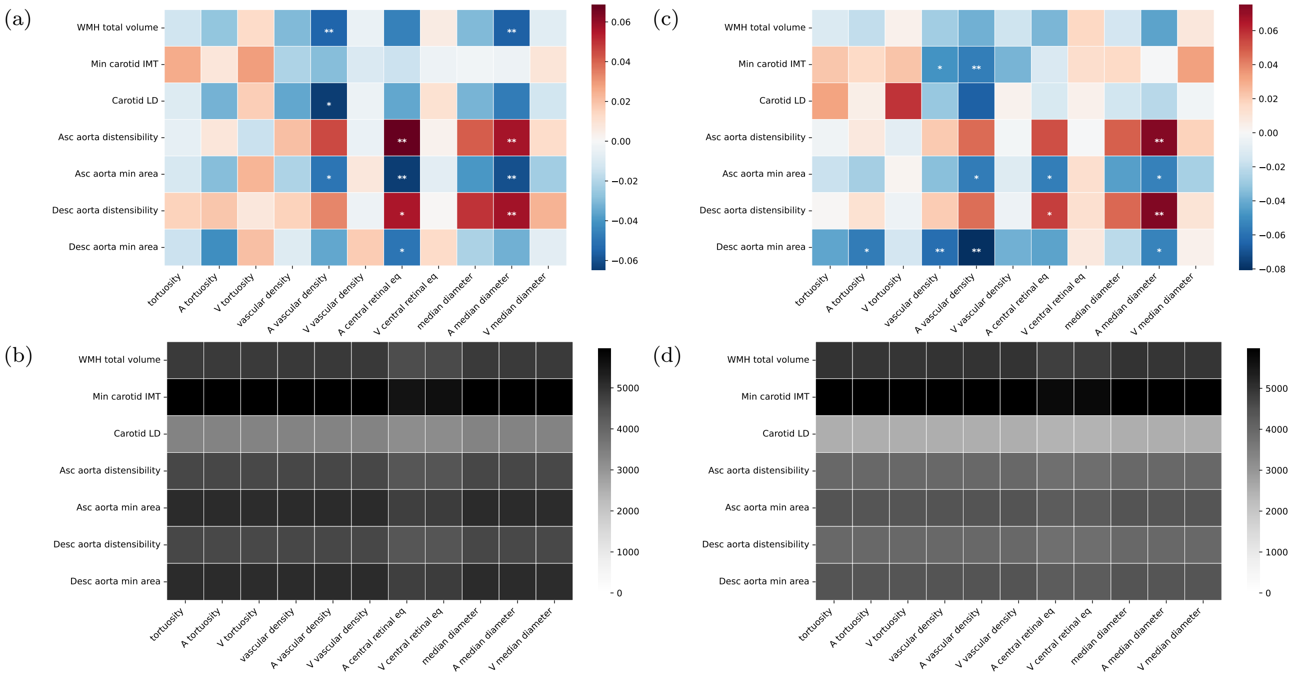


Figure 5: Effect of image-acquisition lag on retinal–systemic correlations. Panels (a)–(b) correspond to the small-gap cohort (left column); panels (c)–(d) to the long-gap cohort (right column).
